# Longitudinal testing for SARS-CoV-2 RNA in day care centers in Hesse, Germany, during increased local incidence and with VOC Alpha as dominant variant: Results of the SAFE KiDS 2 and SAFE KiDS 3 study

**DOI:** 10.1101/2021.06.29.21259633

**Authors:** Barbara Schenk, Sebastian Hoehl, Olga Rudych, Dominic Menger, Samira Farmand, Franziska Wrobel, Emilie Kreutzer, Daniela Gebert, Melanie Flohr, Franziska Berger, Vanessa Weigel, Marhild Kortenbusch, Annemarie Berger, Sandra Ciesek

**Affiliations:** Institute of Medical Virology, University Hospital Frankfurt, Goethe University Frankfurt am Main, Germany, Paul-Ehrlich-Str. 40, D-60596 Frankfurt am Main, Germany

## Abstract

In the summer of 2020, we investigated the rate of inapparent shedding of SARS-CoV-2 in a representative sample of day care centers from Hesse, Germany, and found a low positivity rate during a period of low local community spread.

To investigate the influence of a high local incidence setting, we conducted the *SAFE KiDS 2* study. 577 children and 334 staff members of 47 day care centers were tested for respiratory and gastrointestinal shedding of SARS-CoV-2, and three infections with SARS-CoV-2 in the infectious period were detected. We conclude that viral shedding occurred infrequently while the original “wild-type” variant was dominant.

The more transmissible SARS-CoV-2 variant Alpha (B.1.1.7) became the dominant strain after *SAFE KiDS 2* was concluded. The *SAFE KiDS 3* study investigated the impact of the Alpha variant of SARS-CoV-2 on inapparent viral shedding in the day care setting. In this study, 756 children and 226 staff members from 46 day care centers provided self-collected saliva swabs, the so-called “Lollipop” swabs, which were tested by RT-PCR. In the four-week study period, none of the participants tested positive for SARS-CoV-2 RNA, demonstrating that inapparent shedding of SARS-CoV-2 in the day care setting was also rare during the dominance of the Alpha variant.

The influence of the variant of concern Delta on day care centers has yet to be studied.

## Introduction

In the early pandemic of COVID-19, many countries closed day care centers as measure to curb community spread of the virus. While children of kindergarten age rarely have severe disease when infected with SARS-CoV-2 ^1–3^ measures of public health, such as distancing and wearing of masks, cannot be applied to the same extend in day care centers as in other educational facilities. Nevertheless, a growing body of evidence suggested a limited role of day care centers in community spread of *wild type* SARS-CoV-2 in Central Europe in 2020 and early 2021^4–8^. However, some of these studies were conducted during low community activity^4,5^, limiting the transferability to a setting with increased community spread. One of these studies was the *SAFE KiDS study*, which we conducted in the summer of 2020 in 50 day care centers in the state of Hesse, Germany^4^. In this 12-week longitudinal study, we neither detected respiratory nor gastrointestinal shedding of SARS-CoV-2 in any of 825 children attending day care, and only in two staff members.

But the arrival of the more transmissible variant of concern (VOC) Alpha (B.1.1.7) in Germany in the beginning of 2021^9^ had the potential of new transmission dynamics, which may also influence inapparent viral shedding in day care centers^10,11^. This could also have an impact on effective hygiene concepts of regular operation “under hygienic conditions”.

To evaluate potential changes to the epidemiology of SARS-CoV-2 in the day care setting we investigated the frequency of shedding of SARS CoV-2 in day care centers in Hesse, Germany, in a second and third round of the *SAFE KiDS study: SAFE KiDS 2 and SAFE KiDS 3*, which were conducted from January 18^th^ to February 11^th^, 2021 at a high incidence setting (local 7-day incidence: 23.75 to 281.43 cases / 100,000 inhabitants) during the dominance of the original *wild type* and from May 17^th^ to June 11^th^, 2021 with the Alpha variant being the dominant SARS-CoV-2 variant (local 7-day incidence: 4.73 to 124.61 cases / 100,000 inhabitants) in Germany, respectively. In both studies, children and staff members self-collected swabs at home that subsequently were tested by real time (RT)-PCR for SARS CoV-2.

## Methods

### Study design

The *SAFE KiDS 2* and *SAFE KiDS 3* study in general replicated the study protocol from the *SAFE KiDS* study ^*4*^, with some alterations.

Day care centers that had already participated in the original *SAFE KiDS* study were invited to participate, and, in case the facility declined to participate, further day care centers were chosen from a representative sample and invited. For *SAFE KiDS 2* and *SAFE KiDS 3* study, 47 and 46 facilities were recruited, respectively. 30 participants from each facility, compromising both children and staff members, were invited. Parents were asked to collect swabs weekly from their children at home before visiting the day care center: both a buccal mucosa swab as well as an anal swab (“dual swab”) in the *SAFE KiDS 2* study (according to the original study protocol from the *SAFE KiDS* study) and only a so-called “Lollipop” swab in the *SAFE KiDS 3* study.

To collect the “Lollipop” swab ^12^, children were asked to suck a dry swab for 30 seconds under the supervision of their parents until the swab was saturated with saliva. Staff members provided the same sort of swabs.

Participants received written instructions explaining the goal of the study as well as the swabbing procedure. Written consent was obtained. Testing of children by the parents was only to be performed in compliance with the child, and never forced. Providing samples was voluntary each week for all participants.

### Laboratory testing

Testing for SARS-CoV-2 was performed at the Institute of Medical Virology, Goethe University Frankfurt, Germany. Before proceeding to Real-time (RT)-PCR testing, samples were pooled in a 10-sample group-testing mini-pool protocol, to preserve reagents while maintain high diagnostic sensitivity ^13^. For this purpose, the swabs were incubated and agitated in 1.8 ml virus deactivating buffer for approximately three minutes to generate the archive tubes. After incubation, swabs were squeezed and 10 swabs were pooled into one vial containing 2.7 ml virus deactivating buffer, agitated and incubated for at least three minutes before being removed and discarded. The archive tubes were stored at 4°C and the PCR pool was tested by RT-PCR for SARS-CoV-2 on the Roche cobas® 6800 instrument (Roche diagnostics, Basel, Switzerland) according to manufacturer instructions. In case of a negative result in the pooled sample, all individual samples received a negative test result. When at least one of the two PCR targets (E-gene or ORF-region) was detected, all samples of the pool, the archive tubes, were individually tested. For all individual samples yielding a positive result for either one or both PCR targets, the public health authority was informed in accordance with the German Infection Protection Act.

All positive samples were also screened for the presence of key mutations of SARS-CoV-2 VOC Alpha (N501Y, del69/70, E484K) by melt curve analysis (VirSNiP SARS-CoV-2 Spike N501Y, del69/70, E484K assays; TIB Molbiol, Berlin, Germany), or, if this was not possible due to low viral load, samples were tested with a RT-PCR protocol targeting the S-gene (TaqPathTM COVID-19 CE-IVD RT-PCR Kit, ThermoFischer Scientic, Waltham, USA) to determine whether a drop-out of S-gene detectability occurs in the positive samples of this study, which would be consistent with the presence of the Alpha variant (B.1.1.7) ^14^.

### Statistical analysis

The studies were analyzed in a descriptive statistical assessment due to the low incidence rates.

### Ethical approval

This study protocol was approved by the ethics board of the University Hospital Frankfurt, Goethe University Frankfurt am Main, Germany.

## Results

### Results of the SAFE KiDS 2 study

#### Study participants and sample distribution

47 day care centers participated in this four-week-study. 577 children (age range 5 months to 6 years of age; age median 4 years) and 334 staff members were enrolled and provided at least one swab. 5,019 swabs (2,837 buccal mucosa (56.5 %) and 2,182 anal swabs (43.5 %)) were tested in total. 837 swabs (36.6 %) were provided by staff and 3,182 swabs (63.4 %) were provided by children.

#### Results of testing for SARS-CoV-2 by RT-PCR

We detected SARS-CoV-2 in buccal mucosa swabs of two of 577 children. Gastrointestinal shedding was detected in five additional children, resulting in at least one positive swab in 1.21% of the children (seven out of 577). There was no case with both a positive buccal mucosa and anal swab (table 1). In one out of 334 staff members gastrointestinal shedding of SARS-CoV-2 RNA was detected. All other swabs provided by staff members were negative.

**Table 1.**

| No. | Study week | Day care center | Type of shedding | Child or adult | Ct ORF-1<sup>1)</sup> | Ct E-gene<sup>1)</sup> | Ct S-gene<sup>2)</sup> | Symptoms at time of testing | Confirmation by independent testing of respiratory sample | Associated cases in the day care center | Associated cases in the household | Local-7-day incidence (out of 100,000 inhabitants) |
| --- | --- | --- | --- | --- | --- | --- | --- | --- | --- | --- | --- | --- |
| 1 | 1 | 1 | respiratory | child | 32.57 | 33.53 | 38.38 | none | yes | case no. 2 | Yes | 127.78 |
| 2 | 1 | 1 | gastrointestinal | child | 35.92 | 36.45 | id | none | yes | case no. 1 | Yes | 127.78 |
| 3 | 1 | 2 | respiratory | child | 31.5 | 32.4 | 32.08 | not known | yes | case no. 4 | Yes | 116.85 |
| 4 | 1 | 2 | gastrointestinal | adult | 36.73 | - | 34.51 | fatigue | no | case no. 3 | None | 116.85 |
| 5 | 1 | 3 | gastrointestinal | child | 33.19 | 34.47 | 33.34 | none | Known past infection | None | Yes (prior to study) | 138.71 |
| 6 | 1 | 4 | gastrointestinal | child | 33.85 | 34.33 | 33.27 | none | no | None | Yes (prior to study) | 78.47 |
| 7 | 3 | 5 | gastrointestinal | child | 34.46 | 35.05 | 35.09 | none | no | Yes (prior to study) | Yes (prior to study) | 66.01 |
| 8 | 4 | 6 | gastrointestinal | child | 36.50 | - | id | none | no | None | None | 67.1 |

Characterization of all eight positive SARS-CoV-2 RT-PCR results determined in the *SAFE KiDS 2* study.
<sup>1)</sup> Roche Cobas® SARS-CoV-2 on the Cobas® 6800, Roche diagnostics, Basel, Switzerland
<sup>2)</sup> TaqPath™ COVID-19 CE-IVD RT-PCR Kit, ThermoFischer Scientific, Waltham, USA
id: indeterminable

#### Description of the detected cases

The Ct values of the eight SARS-CoV-2 positive swabs ranged from 31.5 to 36.73 (see table 1). In the first study week of the *SAFE KiDS 2* study, two SARS-CoV-2 infections were detected in children via the buccal mucosa swab and four SARS-CoV-2 infections were detected in three additional children and one staff member via anal swab (see table 1). The SARS-CoV-2 cases occurred in five different day care centers, with the first and second cases and the third and fourth cases detected in study participants of the same day care center. In the second week of the study, no additional SARS-CoV-2 infections were identified with similar numbers of participants. One infection was detected in a child via anal swab in each of the third and fourth study week. All study participants who tested positive had always provided both swab specimens on the day of PCR testing, which were also tested individually by PCR. In no case were both swabs positive.

The eight results obtained were forwarded to the relevant health authorities. Only in the first three cases the detected SARS-CoV-2 infection was confirmed by independent testing of a respiratory swab by the health department. In one case of gastrointestinal viral shedding, a past infection was known; in three cases a past infection was suspected due to SARS-CoV-2 infections in the household of the study participant prior to the study. In none of the eight cases, according to the health authorities, were further transmissions detected in the day care center during the study period (see Table 1). However, the local health authorities determined that in six of the eight cases other persons in the household were also infected with SARS-CoV-2. Only in two cases no other SARS-CoV-2 infections were detected in the household.

#### Community activity of SARS-CoV-2 in Hesse during the study

Local 7-day incidence rates per 100,000 population ranged from 66.01 to 138.71 in the five counties of study participants who tested positive for SARS-CoV-2. Overall, 7-day incidence rates for all participating urban and rural counties ranged from 23.75 to 281.43.

#### Mutation analysis

Due to low viral load in all samples with a positive result for SARS-CoV-2, key mutations from the “variants of concern” Alpha could not be successfully examined using a melt curve analysis. In an alternative approach, we applied the S-gene-targeted PCR assay to test for a drop-out of S-gene-detectability characteristic for the Alpha variant. Due to the high Ct values of the samples, the assay was indeterminable for two samples, for six samples a S-gene drop-out was not detected (meaning the S-gene was detected) (table 1), suggesting that the Alpha variant was not present in these samples.

### Results of the SAFE KiDS 3 study

#### Study participants and sample distribution

The *SAFE KiDS 3* study was conducted from May 17^th^ to June 11^th^, 2021, when the Alpha variant was the dominant virus variant in Hesse, Germany^15^. In the four-week study, 756 children (age range 5 months to 7 years of age; age median 4 years) and 226 staff members of 46 day care centers in Hesse, Germany, participated. In total, 2,964 “Lollipop” swabs were tested for SARS-CoV-2 RNA by RT-PCR. 626 (21.1 %) swabs were provided by staff members and 2,338 (78.9 %) swabs were provided by children.

#### Community activity of SARS-CoV-2 in Hesse during the study

7-day incidence rates for all participating urban and rural counties ranged from 4.73 to 124.61 cases / 100,000 inhabitants.

#### Results of testing for SARS-CoV-2 by RT-PCR

None of the 2,964 swabs tested positive for SARS-CoV-2 RNA. None of the study participants reported that SARS-CoV-2 infection was detected outside of the study.

## Discussion

Building on the *SAFE KiDS* study, we examined the rate of inapparent viral shedding of SARS-CoV-2 in the day care setting during two additional time periods.

In the *SAFE KiDS 2* study, conducted with the “dual swabs” method of testing for both respiratory and gastrointestinal shedding during moderately high local incidence, SARS-CoV-2 was detected in seven out of 577 children (1.2 %) and in one out of 334 staff members (0.3%). Only three of these cases had evidence of respiratory viral shedding. Out of these, SARS-CoV-2 was detected in the buccal mucosa swab collected in the study in two children. In one case, SARS-CoV-2 was detected in an anal swab with subsequent independent testing by the local health department.

In five cases (four in children and one in staff), only gastrointestinal shedding of SARS-CoV-2 was detected, but not concurrent respiratory shedding. This suggests that residual gastrointestinal shedding was present after the acute phase of infection. In fact, in one case, a past infection was known before the beginning of the study; in three other cases, a past infection from household exposure was suspected. The other cases of gastrointestinal viral shedding may also be due to past infections that went unnoticed. Therefore, a low rate of inapparent viral shedding among children attending the day care center can be noted.

To determine the presence of the variant Alpha, which was demonstrated to be more infectious^16,17^, in the *SAFE KiDS 2* study we further analyzed the eight positive samples for an S-gene drop-out by RT-PCR, which would indicate that the Alpha variant was present. No S-gene drop-out was detected. Thus, the effects of the Alpha variant on day care centers have likely not yet influenced the *SAFE KiDS 2* study and were therefore investigated in a third round of the study.

The *SAFE KiDS 3* study was conducted when the Alpha variant was the main circulating variant in Germany ^15^. Since the gastrointestinal detection of SARS-CoV-2 RNA in the *SAFE KiDS 2* study did not provide evidence for an infection during the infectious period, but rather a past infection in the majority of cases, screening for gastrointestinal shedding was omitted in the *SAFE KiDS 3* study. Instead, the so-called “Lollipop” swab was applied to test for respiratory shedding.

Although even more children participated in the *SAFE KiDS 3* study compared to *SAFE KiDS 2* (756 vs. 577), none of the provided swabs tested positive for SARS-CoV-2 in the RT-PCR pooling protocol.

The evaluation of a questionnaire at the end of both studies did not identify a case where our protocol with “dual swab” testing or “Lollipop” testing failed to detect an infection.

In conclusion, interpreted with other recent studies from central Europe ^6,7^ the results of the *SAFE KiDS 2* study suggest a limited role of day care centers in the transmission dynamics of *SARS-CoV-2* even at a high incidence setting. Although the number of infections and transmissions in young children and day care centers has increased upon the arrival of the more contagious variant Alpha in Germany ^10,11^, we did not identify a SARS-CoV-2 infection in the 756 participating children during a period at which Alpha was the dominant circulating variant in the *SAFE KiDS 3* study. The absence of detected SARS-CoV-2 infections in the *SAFE KiDS 3* study is in line with a decline in the local 7-day incidence at the end of the “third wave” in Germany. Seasonality may also have contributed to the decline^18^, in addition to the progressing vaccination campaign. Moreover, a high vaccination quote among the day care staff, as has been reported by the participants of the study, might contribute to decreasing infection numbers in day care centers in the pandemic.

Our study has limitations. This includes that study duration and material was not consistent in all three rounds of the study, limiting comparability of results. All samples were collected without observation by a medical professional, which may have influenced testing sensitivity. Also, since study participation was on a voluntary basis, biases may have also effected the results of the study. We only tested for inapparent shedding, but transmissions were not examined as part of the study.

For the upcoming weeks of the pandemic, it can be assumed that the currently dominant virus variant Alpha will be displaced by the VOC Delta. Whether the even more infectious variant Delta ^19^ will influence transmission among children and thus the incidence of infection in day care centers in the future has to be investigated in further studies.

## Data Availability

The authors confirm that the data supporting the findings of this study are available within the article.

## Figures

## Acknowledgment

We thank Jessica Gille, Regine Jeck, Lena Pompe for technical support and Dr. Ivo Foppa, Hessisches Landesprüfungs- und Untersuchungsamt im Gesundheitswesen, for supplying information on the local incidences throughout Hesse, Germany.

We would further like to thank the Health Protection Authority, City of Frankfurt, and the public health departments of Kreis Bergstraße, Kreis Groß-Gerau and Wetteraukreis for supplying information on the epidemiological context and results of case tracings.

